# Global variation in tests of cognitive and physical function: analysis of six international randomized controlled trials

**DOI:** 10.1101/2023.01.05.22284064

**Authors:** Aristeidis H. Katsanos, Shun Fu Lee, Tali Cukierman-Yaffe, Laura Sherlock, Graciela Muniz-Terrera, Michele Canavan, Raed Joundi, Mukul Sharma, Ashkan Shoamanesh, Andrea Derix, Hertzel C. Gerstein, Salim Yusuf, Martin J. O’Donnell, Jackie Bosch, William N. Whiteley

## Abstract

**Background:** Better understanding of global variation in simple tests of cognition and function would aid the delivery and interpretation of multi-national studies of the prevention of dementia and functional decline.

**Methods:** We aim to describe the variation in simple measures of cognition and function by world region, study, recruitment centre or individual level factors. In six RCTs that measured cognition with the mini-mental state examination (MMSE), Montreal cognitive assessment (MoCA), and instrumental activities of daily living (IADL) with the Standardised Assessment of Everyday Global Activities (SAGEA), we estimated average scores by global region with multilevel mixed-effects models. We estimated the proportion of participants with cognitive or functional impairment with previously defined thresholds (MMSE≤24 or MoCA≤25, SAGEA≥7), and with a country-standardised z-score threshold of cognitive or functional score of ≤-1.

**Results:** In 91,396 participants (mean age 66.6±7.8 years, 31% females) from seven world regions, all global regions differed significantly in estimated cognitive function (z-score differences 0.11–0.45, p<0.001) after accounting for individual-level factors, centre and study. In different regions, the proportion of trial participants with MMSE≤24 or MoCA≤25 ranged from 23–36%; the proportion below a country-standardised z-score threshold of ≤1 ranged from 10–14%. The differences in prevalence of impaired IADL (SAGEA≥7) ranged from 2–6% and by country-standardised thresholds from 3–6%.

**Conclusions:** Accounting for country-level factors reduced large differences between world regions in estimates of cognitive impairment. Measures of IADL were less variable across world regions, and could be used to better estimate dementia incidence in large studies.

**Impact statement:** We certify that this work is novel. After analysing data from a large cohort of participants with a history of cardiovascular disease or cardiovascular risk factors, who were recruited in six international randomised controlled trials (RCTs) we found that accounting for country-level factors reduced large differences between world regions in estimates of cognitive impairment, while measures of functional impairment were less variable across world regions.

**Key Points:**

- Cognitive and functional test scores in randomized controlled clinical trial cohorts vary widely across world regions.
- The difference in cognitive test performance was large in comparison to difference in measures of activities of daily living (ADLs). Accounting for country-level factors reduced the differences between world regions in estimates of cognitive impairment.
- Cognitive test measures were less variable and could be used to better estimate dementia incidence in international studies.

**Why this study matters?:** We found that cognitive and functional test scores in RCT cohorts varied widely across world regions. The difference in cognitive test performance was large in comparison to difference in measures of activities of daily living. The impact of differences on the performance of cognitive tests, which were developed in high-income countries, creates challenges for harmonized studies of cognitive decline prevention in different world regions. Future studies using the same test around the world could standardise cognitive score by country, and consider using in addition measures of instrumental and basic activities of daily living, where there is less variation across world regions.

## Introduction

Although international studies of cardiovascular disease prevention usually measure the incidence of major adverse cardiovascular events, they rarely measure functional impairments, such as cognition or the ability to perform everyday activities.^1, 2^ Although they are important to people, these measures of function are hard to measure and standardize internationally. International standardization of measures of function would be needed for drug approval in each country,^3^ although there are no generally agreed measures.

Culturally appropriate assessments designed to measure cognition or function in individual countries or regions could have internal consistency,^4^ but they are difficult to use at scale or to aggregate between regions. Therefore, using the same assessment tools in all countries is preferable, but requires researchers to recognize and address sources of variation in measuring test performance such as educational attainment, familiarity with testing instruments and the applicability of the functional activities assessed to different regions.^5, 6^ A better understanding of global variation in similar tests of cognition and function, and the sources of variation, would improve measurement, analysis, and interpretation of the results of multi-national studies of cognitive and functional impairment.

We used data from a large cohort of participants with a history of cardiovascular disease or cardiovascular risk factors, in six international randomised controlled trials (RCTs) that assessed antihypertensive, antidiabetic and antithrombotic medications, to describe the variation in simple measures of cognition and function (activities of daily living) by world region. We then sought to determine if variation in the results of cognitive and functional tests between world regions could be explained by study, recruitment centre or individual level factors. We calculated the prevalence of significant cognitive or functional impairment in different world regions, using widely accepted thresholds for cognitive or functional tests, and explored whether country-standardization changed the differences in these estimates between world regions.

## Methods

### Population

We included participants from six large international cardiovascular prevention RCTs.^7-12^ Ongoing Telmisartan Alone and in Combination with Ramipril Global Endpoint Trial (ONTARGET) randomly allocated 25,620 participants with coronary artery disease, peripheral artery disease, cerebrovascular disease, or diabetes mellitus with end-organ damage to ramipril (10 mg once daily), telmisartan (80 mg once daily), or a combination of both.^7^ Telmisartan Randomised AssessmeNt Study in ACE iNtolerant subjects with cardiovascular Disease (TRANSCEND) randomly allocated 5,926 participants meeting the ONTARGET inclusion criteria but intolerant to ACE inhibitors to either telmisartan (80 mg once daily) or placebo.^8^ Outcome Reduction with an Initial Glargine Intervention (ORIGIN) randomly allocated 12,537 participants over the age of 50 with cardiovascular risk factors plus impaired fasting glucose, impaired glucose tolerance, or type 2 diabetes to receive insulin glargine (with a target fasting blood glucose level of 95 mg or less per deciliter [5.3 mmol per liter]) or standard care and to receive n–3 fatty acids or placebo using a 2-by-2 factorial design.^9^ The Cardiovascular Outcomes for People Using Anticoagulation Strategies (COMPASS) trial randomly allocated 27,395 participants with stable atherosclerotic vascular disease to receive rivaroxaban (2.5 mg twice daily) plus aspirin (100 mg once daily), rivaroxaban (5 mg twice daily) monotherapy, or aspirin monotherapy (100 mg once daily).^10^ New Approach Rivaroxaban Inhibition of Factor Xa in a Global Trial versus ASA to Prevent Embolism in Embolic Stroke of Undetermined Source (NAVIGATE ESUS) randomly allocated 7,213 participants with a recent embolic stroke of presumed cardioembolic source to either rivaroxaban (15 mg once daily) or aspirin (100 mg once daily).^11^ The Heart Outcomes Prevention Evaluation (HOPE)–3 trial randomly allocated 12,705 participants with intermediate cardiovascular disease risk to receive rosuvastatin (10 mg once daily) or placebo and to receive a combination of candesartan/ hydrochlorothiazide (16/12.5 mg once daily) or placebo, using a 2-by-2 factorial design.^12^ In HOPE-3 cognitive assessments were obtained only for those 70 years and older.^12^

### Exposures

We grouped countries into seven world regions: North America, Oceania and Western Europe; South America; Eastern Europe and Russia; East Asia; South Asia; Africa; and West Asia (Supplementary Table 1).^13^

Individual participant covariates, measured as continuous variables at either the run-in or randomization visit, were: age in years, body mass index (BMI), waist circumference; systolic and diastolic blood pressure; fasting glucose; and hemoglobin A1c % measured in Diabetes Control and Complications Trial (DCCT) units. We included the following binary variables (yes versus no, excluding missing or unknown) assessed at either the run-in or randomization visit: sex; regular alcohol use (drinking alcohol once or more per week); employment; history of coronary or cerebrovascular disease (transient ischemic attack, stroke, myocardial infarction, angina, heart failure), atrial fibrillation, peripheral arterial disease, hyperlipidemia, hypertension, diabetes mellitus, other non-vascular co-morbidities (cancer, renal dysfunction, liver disease, depression) and history of fall or fracture. Education was categorized as: none or less than primary, primary or secondary, or trade school/college/university. We used the components of the EQ-5D assessment in each trial to derive information on depression and disability.^14^

### Outcomes

For different world regions we presented the results of a cognitive assessment at either the run-in or randomization visit, which was performed using two cognitive tests: the Mini-Mental State Examination (MMSE), a 30-point assessment that addresses seven different cognitive domains (range 0-30, with higher scores meaning better cognition)^15^ and the Montreal Cognitive Assessment (MoCA), a 30-point assessment that addresses seven different cognitive domains (range 0-30, with higher scores meaning better cognition). For the MoCA test an additional point is granted to individuals with 12 or fewer years of education. The MMSE was performed in the ORIGIN, TRANSCEND and ONTARGET trials,^7-9^ while MoCA was performed in HOPE-3, COMPASS and NAVIGATE ESUS.^10-12^ The short version of the MoCA test (maximum score of 12) was completed in HOPE-3, and in our analyses was normalized it to a 30-point scale to allow comparison with other studies.^16, 17^ In a subset of trials, we measured the Standardised Assessment of Everyday Global Activities (SAGEA), a 15-item patient-reported outcome measure developed to measure functional status (basic, instrumental, and cognitive activities of daily living) in patients with vascular disease (range 0-45, with higher scores denoting worse function).^18^ The individual items and scoring for the SAGEA is outlined in the Appendix. All tests were administered in the language of participants.

### Statistical analysis

Participants with cognitive or functional assessment available either at run-in or randomization visits were included in the analyses. To account for differences in the study populations we standardised cognitive and functional test scores to the population of each study (study-standardised) or the country’s population (country-standardised). Each participant’s raw score was subtracted from the study’s or country’s specific mean score, and the difference was then divided by the study’s or country’s specific baseline standard deviation (SD), respectively. Given the comparability in z-scores by age between MMSE and MoCA (Supplementary Fiigure 1),^19^ we pooled the standardised z-scores of these two tests.

We then constructed multilevel mixed-effects models using the study-standardised or country-standardised test scores as the dependent variable, the baseline participant covariates as fixed-effects modifiers and the recruiting centre as a random-effects modifier (allowing testing of between centre heterogeneity). In the unadjusted model we provide the crude estimates; in the age-sex-adjusted model we adjust only for the individual’s age and sex and in the maximally-adjusted model we adjust for the individual’s age, sex, education, BMI, history of cardiovascular disease, history of cardiovascular risk factors, history of non-cardiovascular diseases and baseline blood pressure. We used the North America, Oceania and Western Europe group as reference, since cognitive and functional tests were developed in countries from these geographical regions, and present the unadjusted and adjusted mean differences and corresponding 95% confidence intervals (95%CI) for other world regions.

We estimated the prevalence of cognitive impairment in different world regions with absolute score thresholds for the MMSE (≤24 points) or MoCA (≤25 points),^20, 21^ and with a threshold for the study- and country-standardised scores that corresponds to the thresholds of MMSE (24 points or less) or MoCA (25 points or less) in the North America, Oceania and Western Europe group (z≤1.0).^22^ We then applied this z-score threshold to the other world regions. The same approach was used to estimate the prevalence of functional impairment in different world regions with the absolute threshold of 7 or more points in the SAGEA score,^18^ and then applied the respective z-score threshold for a score of 7 to study- or country-standardised distributions from the other world regions. We calculated the intraclass correlation coefficient (ICC), the proportion of variation within clusters (world regions) over the total variation, for each analysis to estimate the variation of cognitive or functional impairment between the different world regions. The chi-square test was used to test the homogeneity of intraclass correlations.

### Standard Protocol Approvals, Registrations, and Patient Consents

The present work is a post-hoc analysis of previously published randomized controlled clinical trials coordinated by the Population Health Research Institute and was exempt from ethics review board approval. For the collection of data in original studies, informed consent was obtained by primary investigators.

## Results

We analyzed data from 91,396 participants in six RCTs from 55 countries recruited between 2001 and 2017 in seven world regions (Supplementary Table 1): North America, Oceania, and Western Europe (39,668); South America (17,923); Eastern Europe and Russia (13,433); East Asia (11,667); South Asia (5,146); Africa (2,273) and West Asia (1,286). Participants had a mean age of 66.6 years (SD 7.8), and 31% were females. Few participants were smokers (17%) or regularly used alcohol (25%). Most participants were educated to primary or secondary level (54%), 39% to higher technical level and 3% had no or less than primary education. Almost a third of participants were employed at study enrolment, 18% had self-reported depression and 9% had self-reported disability based on the EQ-5D questionnaire assessment. More than 90% of the participants had at least one cardiovascular risk factor and 69% had a history of one or more cardiovascular diseases (Supplementary Table 2). Additional participant and country characteristics are provided in Supplementary Table 3.

Participants’ baseline characteristics demonstrating the greatest variability included alcohol consumption (ranging from 12% in South Asia to 30% in North America, Oceania and Western Europe), self-reported depression (ranging from 12% in Africa to 24% in South America) and self-reported disability (ranging from 3% in East Asia to 18% in South Asia). The prevalence of cardiovascular disease comorbidity at baseline study assessment also varied between the different world regions (Supplementary Table 2).

Study-standardised cognitive test scores were lower in people who were older in all world regions except Africa, with large differences between regions (Figure 1A). However, when cognitive scores were standardised to country, older people had consistently lower cognitive scores in all regions, and the variation in z-scores between regions was attenuated (Figure 1B).

**Figure 1.**
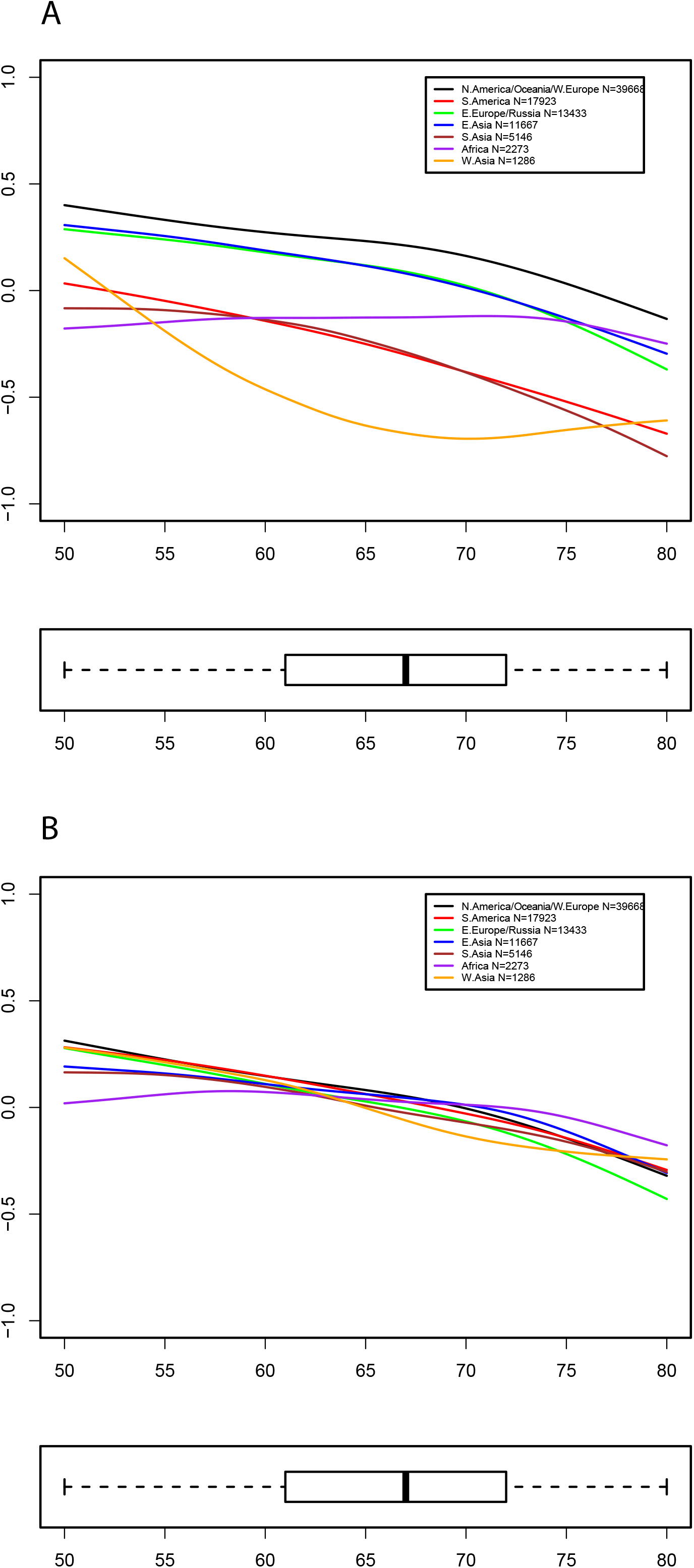
Fitting splines on the association between individual’s age and (A) study-standardised or (B) country-standardised cognitive scores by different world regions. Box plot illustrate the median, interquartile range and range of scores in the whole study population. (STD: standardised; NA: North America; SA: South America; EE: Eastern Europe; EAsia: East Asia; SAsia: South Asia; W.Asia: West Asia)

Cognitive test z-scores standardised for study (adjusted for individual characteristics and centre from which they were recruited), were significantly lower in world regions (p<0.001 for all regions) compared with North America, Oceania and Western Europe (greatest difference for South America z-score mean difference -0.45 [95% CI: -0.51 to -0.40]). Standardizing the cognitive test scores for country (adjusting for individual characteristics and centre) reduced variation between world regions. South America, Eastern Europe and Russia, East Asia, Africa and West Asia had modest differences (all z-score mean difference >-0.1, p>0.05) from North America, Oceania and Western Europe, although the difference was slightly greater for South Asia (z-score mean difference: -0.12, 95%CI: -0.22 to -0.02; Figure 2, Supplementary Table 4).

**Figure 2.**
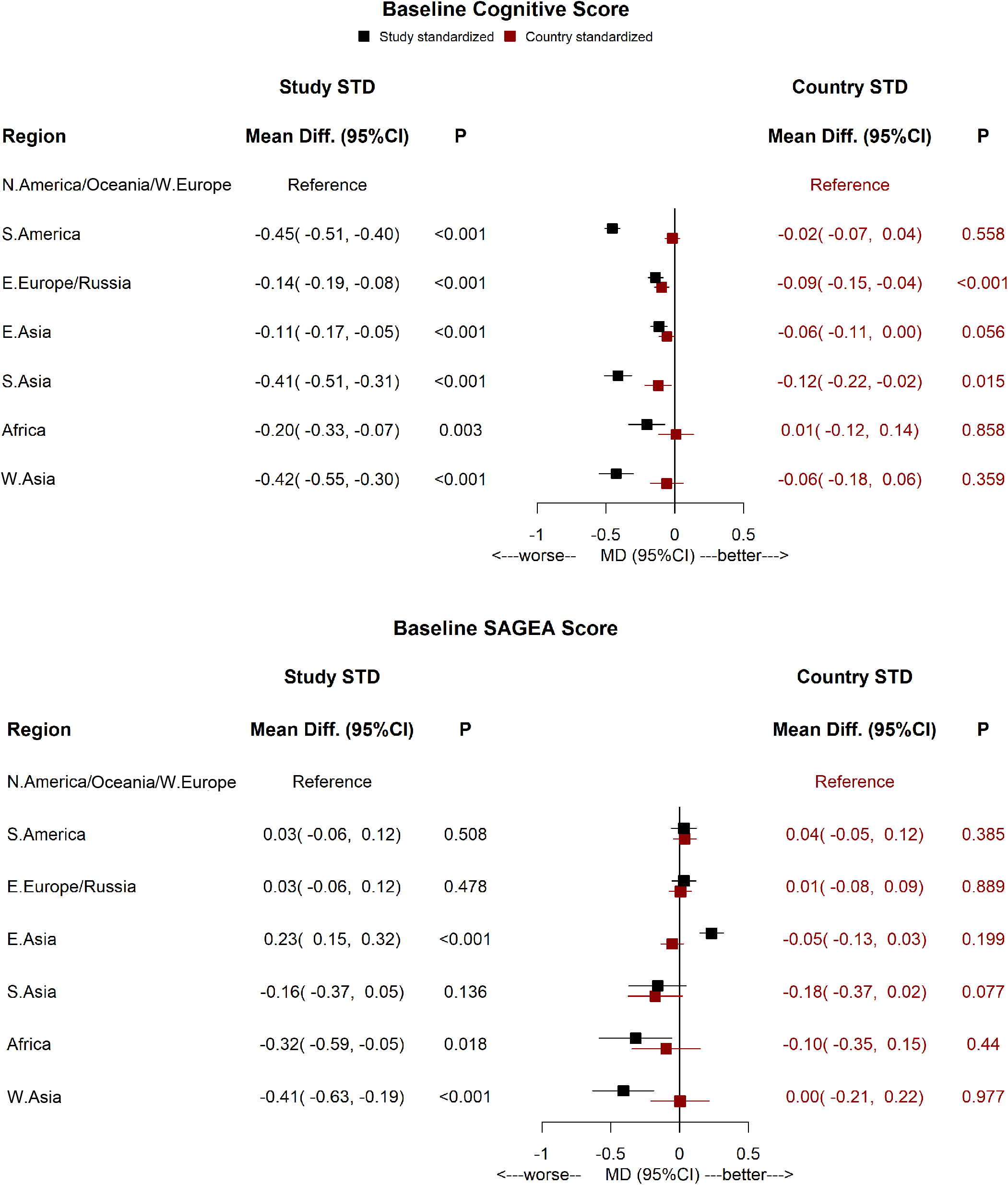
Mean differences in study-standardised or country-standardised baseline cognitive and functional test scores between the North America, Oceania and Western Europe group and other world regions, adjusted for individual participant characteristics and centre of recruitment. (SAGEA: Standardised Assessment of Global activities in the Elderly; NA: North America; SA: South America; EE: Eastern Europe; EAsia: East Asia; SAsia: South Asia; W.Asia: West Asia)

Widely used absolute thresholds for MMSE (≤24 points) or MOCA (≤25 points) classified 27% of the overall population with cognitive impairment, with proportions ranging from 23% in North America, Oceania and Western Europe to 36% in South America and West Asia (ICC=0.032, χ^2^=1787). Using the corresponding absolute values z-score threshold of -1.0 (Supplementary Table 5) in study-standardised (not shown) and country standardized cognitive scores led to a lower proportion of participants in each region classified with cognitive impairment (Figure 3).

**Figure 3.**
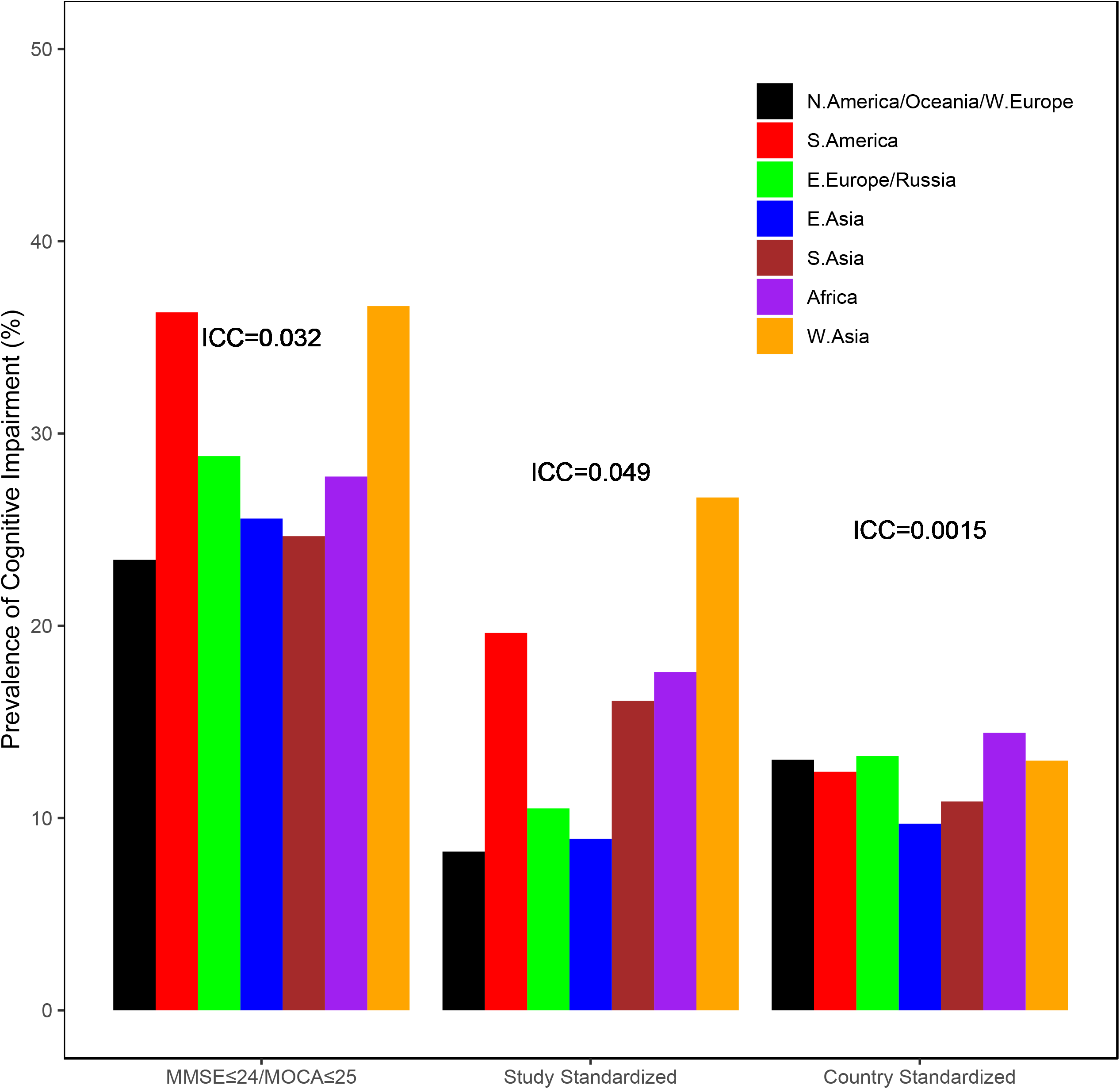
Estimated prevalence of cognitive impairment in different word regions using absolute (MMSE ≤24) or MOCA ≤25) or relative thresholds (z-score≤-1.0) based on the study-standardised or country-standardised mean cognitive test scores. Intraclass correlation coefficient (ICC) is the proportion variance within clusters over the total variance. (NA: North America; SA: South America; EE: Eastern Europe; EAsia: East Asia; SAsia: South Asia; W.Asia: West Asia)

The study-standardized functional SAGEA scores (adjusted for individual characteristics and centre) were lower in Africa (z-score mean difference: -0.32, 95%CI: -0.59 to -0.05) and West Asia (-0.41, 95%CI: -0.63 to -0.19), and higher in East Asia (+0.23, 95%CI: 0.15 to 0.32) compared with North America, Oceania and Western Europe. However, after country standardization (and adjusting for individual characteristics and centre), there were no large or significant differences in SAGEA scores between world regions (all z-score mean differences ≤0.18, p>0.05; Figure 2 & Supplementary Table 4).

With a threshold ≥7 points in SAGEA score, 4% of the population was classified with functional impairment, with prevalence rates between regions from 2 to 7% (ICC=0.028, χ^2^ =89). A z-score of -1.0 in study-standardised distributions and country-standardised distributions resulted in 3% (range 2-6%, ICC=0.024, χ^2^=191) and 4% (range 2-4%, ICC=0.0007, χ^2^=41) prevalence rates of functional impairment, respectively (Figure 4).

**Figure 4.**
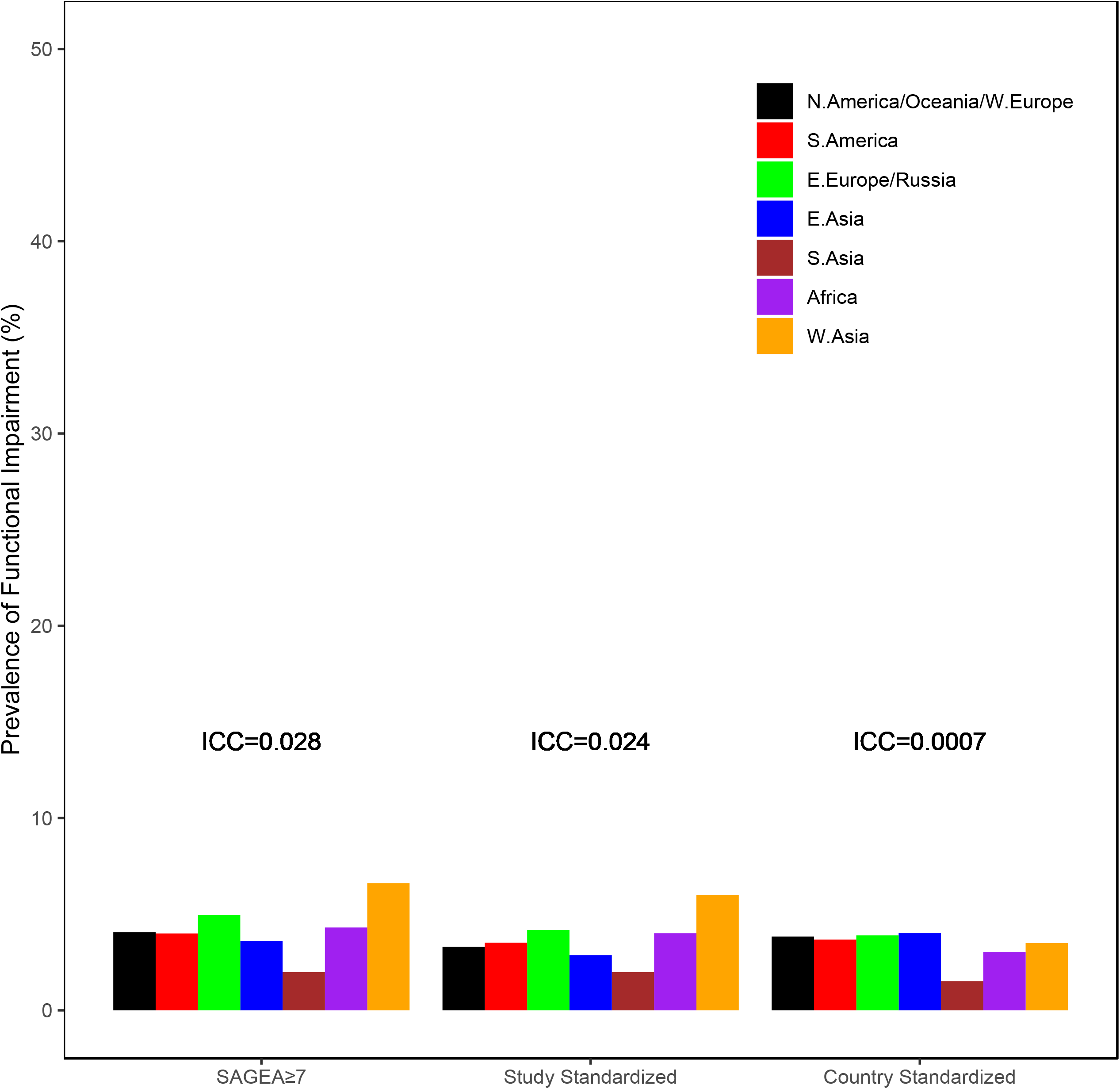
Estimated prevalence of functional impairment in different word regions using absolute (SAGE ≥7) or relative thresholds (z-score≤-1.0) based on the study-standardised or country-standardised mean functional test scores. Intraclass correlation coefficient (ICC) is the proportion variance within clusters over the total variance. (NA: North America; SA: South America; EE: Eastern Europe; EAsia: East Asia; SAsia: South Asia; W.Asia: West Asia)

## Discussion

We found that cognitive and functional test scores in RCT cohorts varied widely across world regions. The difference in cognitive test performance was large in comparison to difference in measures of activities of daily living.

These differences in cognitive test scores attenuated modestly after adjusting for individual level factors, or randomisation centre, a surrogate for the socio-economic status of the area. However, when standardising for country, the differences attenuated substantially, suggesting that unmeasured country-level factors accounted for much of the difference in test performance. Better measurement of educational experience and literacy could further reduce differences in cognitive test performance. Our results are in accordance with previous literature suggesting that there is substantial variation in cognitive test performance between different world regions.^4, 6, 13^ The variability in cognitive test performance between different world regions has previously been associated with modifiable and non-modifiable individual-level factors,^23-27^ and the socio-economic status of the area of residence for an individual.^28-31^

The prevalence of cognitive impairment using widely accepted absolute cognitive score thresholds was very large, suggesting factors related to test performance rather than cognitive impairment were responsible for these differences and questioning the suitability of these thresholds for population-based studies. This problem has been identified previously in population-based studies.^32^ In Sweden, a MoCA threshold of 25 points or less classified 37.3% of the population as cognitively impaired.^33^ A systematic review and meta-analysis of the cross-cultural applicability of MoCA screening for mild cognitive impairment found a wide range of suggested cut-offs both across countries and within regions.^34^ For example, the cut-off scores of the MoCA were different for five Chinese versions, ranging between 23 and 25 for mild cognitive impairment and between 19 and 24 for dementia.^35^

In our study, functional measurement showed less variation by region and demonstrate that the use of measures of activities of daily living along with cognitive scores could be used to support a diagnosis of significant cognitive impairment in international studies. Functional measures showed much less regional variation in scores than measures of cognition even before taking into account individual, centres or country-level factors. Tests that use functional measures to measure impairment, although less sensitive might be more consistent between world regions.

Our study had a number of limitations. We analyzed cognitive and functional assessment scores from participants recruited in international large-scale RCTs of cardiovascular interventions. Although these trials were performed in multiple countries and centres around the world, resulting in a multi-ethnic and culturally diverse population, these participants were not representative of their country populations, and our findings are only relevant for trials population. Individuals who agree to participate in RCTs can be very different from the general population on factors beyond the overt inclusion/exclusion criteria. There were substantial differences in the total number of individuals from each of the world regions, leading to increased uncertainty and less precise fit of statistical models for the regions with fewest participants (i.e. West Asia, Africa). Additionally, the number of countries included in each world region differed, e.g. only one country was included for Africa group and 20 countries were included for North America, Oceania and Western Europe group (Supplementary Table 1). Finally, there was no clinical diagnosis of dementia or mild cognitive impairment, although all patients were able to consent, and none had recorded dementia at baseline.^36^

The impact of differences on the performance of cognitive tests, which were developed in high-income countries, creates challenges for harmonized studies of cognitive decline prevention in different world regions.^37^ Universal cut-offs for cognitive decline may lead to erroneous conclusions about an individual’s cognitive ability and pose barriers in the interpretation and collation of the results from these tests in international studies.^38^ New tests could be developed for each country, although this increases study cost and complexity. Future studies using the same test around the world could standardise cognitive score by country, and consider using in addition measures of instrumental and basic activities of daily living, where there is less variation across world regions.

## Data Availability

All data produced in the present study are available upon reasonable request to the authors

## Notes

**Conflict of Interest Authors** report no conflicts of interest relevant to this particular manuscript

### Competing Interest Statement

The authors have declared no competing interest.

### Funding Statement

This study did not receive any funding

## References

1. Iadecola C, Yaffe K, Biller J, et al; American Heart Association Council on Hypertension; Council on Clinical Cardiology; Council on Cardiovascular Disease in the Young; Council on Cardiovascular and Stroke Nursing; Council on Quality of Care and Outcomes Research; and Stroke Council. Impact of hypertension on cognitive function: a scientific statement from the American Heart Association. Hypertension. 2016;68:e67–e94.

2. Collins R, Reith C, Emberson J, et al. Interpretation of the evidence for the efficacy and safety of statin therapy. Lancet. 2016;388:2532–2561.

3. Katz R. FDA: evidentiary standards for drug development and approval. NeuroRx. 2004;1:307–16.

4. Prince M, Acosta D, Chiu H, Scazufca M, Varghese M; 10/66 Dementia Research Group. Dementia diagnosis in developing countries: a cross-cultural validation study. Lancet. 2003;361:909–17.

5. Max Roser and Esteban Ortiz-Ospina (2016) - “Literacy”. Published online at OurWorldInData.org. Retrieved from: ‘https://ourworldindata.org/literacy’ [Online Resource]

6. Ng KP, Chiew HJ, Lim L, Rosa-Neto P, Kandiah N, Gauthier S. The influence of language and culture on cognitive assessment tools in the diagnosis of early cognitive impairment and dementia. Expert Rev Neurother. 2018;18:859–869.

7. ONTARGET Investigators, Yusuf S, Teo KK, Pogue J, et al. Telmisartan, ramipril, or both in patients at high risk for vascular events. N Engl J Med. 2008;358:1547–59.

8. Telmisartan Randomised AssessmeNt Study in ACE iNtolerant subjects with cardiovascular Disease (TRANSCEND) Investigators, Yusuf S, Teo K, Anderson C, et al. Effects of the angiotensin-receptor blocker telmisartan on cardiovascular events in high-risk patients intolerant to angiotensin-converting enzyme inhibitors: a randomised controlled trial. Lancet. 2008;372:1174–83.

9. ORIGIN Trial Investigators, Gerstein HC, Bosch J, Dagenais GR, et al. Basal insulin and cardiovascular and other outcomes in dysglycemia. Basal insulin and cardiovascular and other outcomes in dysglycemia. N Engl J Med. 2012;367:319–328.

10. Eikelboom JW, Connolly SJ, Bosch J, et al. Rivaroxaban with or without aspirin in stable cardiovascular disease. N Engl J Med. 2017;377:1319–1330.

11. Hart RG, Sharma M, Mundl H, et al. Rivaroxaban for Stroke Prevention after Embolic Stroke of Undetermined Source. N Engl J Med. 2018;378:2191–2201.

12. Yusuf S, Lonn E, Pais P, et al. Blood-Pressure and Cholesterol Lowering in Persons without Cardiovascular Disease. N Engl J Med. 2016;374:2032–2043.

13. Kalaria RN, Maestre GE, Arizaga R, et al. Alzheimer’s disease and vascular dementia in developing countries: prevalence, management, and risk factors. Lancet Neurol. 2008;7:812–26.

14. EuroQol Group. EuroQol--a new facility for the measurement of health-related quality of life. Health Policy. 1990;16:199–208.

15. Folstein MF, Folstein SE, McHugh PR. “Mini-mental state”. A practical method for grading the cognitive state of patients for the clinician. J Psychiatr Res. 1975;12:189–98.

16. Nasreddine ZS, Phillips NA, Bédirian V, et al. The Montreal Cognitive Assessment, MoCA: a brief screening tool for mild cognitive impairment. J Am Geriatr Soc. 2005;53:695–9.

17. Dong Y, Koay WI, Yeo LL, Chen CL, Xu J, Seet RC, Lim EC. Rapid Screening for Cognitive Impairment in Parkinson’s Disease: A Pilot Study. Parkinsons Dis. 2015;2015:348063.

18. Spence J, Bosch J, Chongsi E, et al. Standardized Assessment of Global activities in the Elderly scale in adult cardiac surgery patients. Br J Anaesth. 2021;127:539–546.

19. Yang H, Yim D, Park MH. Converting from the Montreal Cognitive Assessment to the Mini-Mental State Examination-2. PLoS One. 2021;16:e0254055.

20. Creavin ST, Wisniewski S, Noel-Storr AH, et al. Mini-Mental State Examination (MMSE) for the detection of dementia in clinically unevaluated people aged 65 and over in community and primary care populations. Cochrane Database Syst Rev. 2016:CD011145.

21. Davis DH, Creavin ST, Yip JL, Noel-Storr AH, Brayne C, Cullum S. Montreal Cognitive Assessment for the detection of dementia. Cochrane Database Syst Rev. 2021;7:CD010775.

22. Perkins NJ, Schisterman EF. The inconsistency of “optimal” cutpoints obtained using two criteria based on the receiver operating characteristic curve. Am J Epidemiol. 2006;163:670–5.

23. Livingston G, Huntley J, Sommerlad A, et al. Dementia prevention, intervention, and care: 2020 report of the Lancet Commission. Lancet. 2020;396:413–446.

24. Daviglus ML, Bell CC, Berrettini W, et al. National Institutes of Health State-of-the-Science Conference statement: preventing alzheimer disease and cognitive decline. Ann Intern Med. 2010;153:176–81.

25. Kivimäki M, Singh-Manoux A, Pentti J, et al. Physical inactivity, cardiometabolic disease, and risk of dementia: an individual-participant meta-analysis. BMJ. 2019;365:l1495.

26. Albanese E, Lombardo FL, Prince MJ, Stewart R. Dementia and lower blood pressure in Latin America, India, and China: a 10/66 cross-cohort study. Neurology. 2013;81:228–35.

27. Ott A, Breteler MM, van Harskamp F, et al. Prevalence of Alzheimer’s disease and vascular dementia: association with education. The Rotterdam study. BMJ. 1995;310:970–3.

28. Selvamani Y, Arokiasamy P. Association of life course socioeconomic status and adult height with cognitive functioning of older adults in India and China. BMC Geriatr. 2021;21:354.

29. Yu X, Zhang W, Kobayashi LC. Duration of Poverty and Subsequent Cognitive Function and Decline Among Older Adults in China, 2005-2018. Neurology. 2021;97:e739–e746.

30. Paradela RS, Ferreira NV, Nucci MP, et al. Relation of a Socioeconomic Index with Cognitive Function and Neuroimaging in Hypertensive Individuals. J Alzheimers Dis. 2021;82:815–826.

31. Okamoto S. Socioeconomic factors and the risk of cognitive decline among the elderly population in Japan. Int J Geriatr Psychiatry. 2019;34:265–271

32. Borland E, Nägga K, Nilsson PM, Minthon L, Nilsson ED, Palmqvist S. The Montreal Cognitive Assessment: Normative Data from a Large Swedish Population-Based Cohort. J Alzheimers Dis. 2017;59:893–901.

33. Chandra V, Ganguli M, Ratcliff G, et al. Studies of the epidemiology of dementia: comparisons between developed and developing countries. Aging Milano 1994; 6: 307–21.

34. O’Driscoll C, Shaikh M. Cross-Cultural Applicability of the Montreal Cognitive Assessment (MoCA): A Systematic Review. J Alzheimers Dis. 2017;58:789–801.

35. Tan JP, Li N, Gao J, et al. Optimal cutoff scores for dementia and mild cognitive impairment of the Montreal Cognitive Assessment among elderly and oldest-old Chinese population. J Alzheimers Dis. 2015;43:1403–12.

36. Llibre Rodriguez JJ, Ferri CP, Acosta D, et al. Prevalence of dementia in Latin America, India, and China: a population-based cross-sectional survey. Lancet. 2008;372:464–474.

37. Magklara E, Stephan BCM, Robinson L. Current approaches to dementia screening and case finding in low- and middle-income countries: Research update and recommendations. Int J Geriatr Psychiatry. 2019;34:3–7.

38. Whiteley WN, Anand S, Bangdiwala SI, et al. Are large simple trials for dementia prevention possible? Age Ageing. 2020;49:154–160.

