## Appendix for "Global variation in tests of cognitive and physical function: analysis of six international randomized controlled trials"

**Supplementary Table 1.** Overview of participant numbers per study and country, as grouped into different world regions.

| **Country/ study** | COMPASS | HOPE-3 | NAVIGATE ESUS | ONTARGET/ TRANSCEND | ORIGIN |
| --- | --- | --- | --- | --- | --- |
| North America, Oceania, Western Europe | | | | | |
| Canada | 2443 | 1156 | 573 | 2945 | 983 |
| Denmark | 575 | . | 146 | 487 | 362 |
| Finland | 119 | . | 131 | 374 | 221 |
| France | 192 | . | 288 | 841 | 81 |
| Germany | 766 | . | 198 | 1851 | 410 |
| Ireland | 382 | . | 13 | 182 | 263 |
| Italy | 1013 | . | 334 | 1085 | 170 |
| Netherlands | 2522 | 117 | . | 1201 | 424 |
| Sweden | 735 | 117 | 71 | 548 | 463 |
| Switzerland | 58 | . | 125 | 229 | 28 |
| United Kingdom | 541 | 69 | 435 | 976 | 109 |
| USA | 1475 | . | 345 | 3306 | 318 |
| Australia | 353 | 45 | 119 | 1374 | 202 |
| Belgium | 455 | . | 90 | 908 | . |
| Austria | . | . | 174 | 168 | 103 |
| Portugal | . | . | 154 | 211 | . |
| Spain | . | . | 618 | 594 | 138 |
| Norway | . | . | . | 240 | 159 |
| Bermuda | . | . | . | . | 13 |
| New Zealand | . | . | . | 754 | . |
| South America |  |  |  |  |  |
| Argentina | 2789 | 1459 | 265 | 1662 | 2222 |
| Brazil | 1515 | 551 | 179 | 1156 | 413 |
| Chile | 641 | . | 194 | . | 168 |
| Colombia | 942 | 1463 | . | . | 708 |
| Ecuador | 257 | 397 | . | . | . |
| Mexico | . | . | 108 | 492 | 211 |
| Venezuela | . | . | . | . | 131 |
| Eastern Europe (including Russia) | | | | | |
| Czech Republic | 1553 | 70 | 243 | 617 | . |
| Hungary | 734 | 263 | 224 | 661 | 292 |
| Poland | 518 | . | 284 | 919 | 187 |
| Russia | 682 | 190 | 328 | 625 | 524 |
| Slovakia | 92 | 10 | . | 407 | 37 |
| Ukraine | 821 | 333 | . | 817 | . |
| Romania | 423 | . | . | . | 846 |
| Greece | . | . | 76 | 266 | . |
| Belarus | . | . | . | . | 127 |
| Croatia | . | . | . | . | 115 |
| Estonia | . | . | . | . | 47 |
| Latvia | . | . | . | . | 54 |
| Lithuania | . | . | . | . | 48 |
| East Asia |  |  |  |  |  |
| China | 1086 | 3677 | 491 | 1679 | 454 |
| Japan | 1556 | . | 647 | . | . |
| South Korea | 415 | 14 | 212 | 416 | 131 |
| Hong Kong | . | . | . | 401 | . |
| Taiwan | . | . | . | 488 | . |
| South Asia | | | | | |
| India | . | 1824 | . | . | 390 |
| Malaysia | 247 | 87 | . | 267 | . |
| Philippines | 651 | 571 | . | 424 | 133 |
| Singapore | . | . | . | 93 | . |
| Thailand | . | . | . | 459 | . |
| Africa |  |  |  |  |  |
| South Africa | 581 | 211 | 49 | 832 | 600 |
| West Asia |  |  |  |  |  |
| Israel | 263 | 81 | 60 | . | 146 |
| Turkey | . | . | 39 | 247 | 106 |
| UAE | . | . | . | 344 | . |

COMPASS: Cardiovascular Outcomes for People Using Anticoagulation Stratgies; HOPE-3: Heart Outcomes Prevention Evaluation-3; NAVIGATE ESUS: New Approach Rivaroxaban Inhibition of Factor Xa in a Global Trial versus ASA to Prevent Embolism in Embolic Stroke of Undetermined Source; ORIGIN: Outcome Reduction With Initial Glargine Intervention; ONTARGET: ONgoing Telmisartan Alone and in combination with Ramipril Global Endpoint Trial; TRANSCEND: Telmisartan Randomized AssessmeNt Study in ACE iNtolerant subjects with cardiovascular Disease

**Supplementary Table 2.** Study participant characteristics in different world regions

| **Description** | **North America, Oceania, Western Europe** | **South America** | **Eastern Europe**  **and Russia** | **East Asia** | **South Asia** | **Africa** | **West Asia** |
| --- | --- | --- | --- | --- | --- | --- | --- |
| Number of participants, N | 39668 | 17923 | 13433 | 11667 | 5146 | 2273 | 1286 |
| Years recruited | 2001-2017 | 2002-2017 | 2002-2017 | 2002-2017 | 2002-2016 | 2002-2017 | 2002-2017 |
| Years of follow-up, median (IQR) | 4.41 (1.97-5.02) | 4.44 (1.80-5.83) | 3.84 (1.61-5.00) | 4.48 (2.01-5.34) | 5.00 (4.09-5.59) | 4.83 (2.57-5.79) | 4.51 (2.79-5.29) |
| Baseline Characteristics |  |  |  |  |  |  |  |
| Age, years, mean (SD) | 66.97 (7.73) | 66.33 (7.85) | 65.99 (7.96) | 66.25 (7.82) | 66.39 (7.11) | 66.71 (8.53) | 65.42 (7.23) |
| BMI, mean (SD) | 28.51 (4.84) | 28.65 (5.11) | 28.39 (4.73) | 26.66 (4.1) | 26.89 (4.74) | 28.71 (4.78) | 28.8 (4.62) |
| Waist circumference, cm, mean (SD) | 98.89 (13.56) | 98.77 (13.46) | 98.85 (12.76) | 94.16 (11.24) | 93.95 (12.45) | 98.58 (12.49) | 96.98 (12.86) |
| Female, N (%) | 11282 (28.44%) | 5667 (31.62%) | 4134 (30.77%) | 3942 (33.79%) | 2179 (42.34%) | 777 (34.18%) | 454 (35.33%) |
| Current smoker, N (%) | 5988 (15.11%) | 3037 (16.95%) | 2522 (18.79%) | 2498 (21.41%) | 790 (15.36%) | 281 (12.36%) | 208 (16.19%) |
| Regular alcohol, N (%) | 10772 (30.11%) | 3922 (22.93%) | 2765 (22.58%) | 2055 (19.99%) | 601 (11.69%) | 594 (26.83%) | 265 (22.34%) |
| Education |  |  |  |  |  |  |  |
| -Less than primary or none, N (%) | 629 (1.6%) | 867 (4.92%) | 310 (2.33%) | 758 (6.58%) | 287 (5.68%) | 41 (1.82%) | 9 (0.71%) |
| -Primary/Secondary, N (%) | 19892 (50.64%) | 11165 (63.4%) | 6468 (48.7%) | 6228 (54.1%) | 2956 (58.52%) | 1132 (50.38%) | 563 (44.19%) |
| -Trade school/College/University, N (%) | 16966 (43.19%) | 5218 (29.63%) | 5846 (44.02%) | 3671 (31.89%) | 1808 (35.79%) | 1043 (46.42%) | 634 (49.76%) |
| Socieconomic |  |  |  |  |  |  |  |
| Employment, N (%) | 5433 (31%) | 4496 (32.6%) | 1973 (24.87%) | 2186 (29.94%) | 1042 (26.74%) | 343 (24.8%) | 133 (22.35%) |
| Depression, N (%) | 6287 (15.85%) | 4234 (23.62%) | 2706 (20.14%) | 1686 (14.45%) | 930 (18.07%) | 284 (12.49%) | 238 (18.51%) |
| Disability, N (%) | 1609 (8.14%) | 870 (12.12%) | 359 (6.94%) | 177 (2.65%) | 658 (17.66%) | 120 (11.51%) | 63 (9.38%) |
| Cardiovascular risk factors |  |  |  |  |  |  |  |
| - 0 risk factor, N (%) | 4085 (10.3%) | 1694 (9.45%) | 931 (6.93%) | 1175 (10.07%) | 507 (9.85%) | 143 (6.29%) | 77 (5.99%) |
| - 1 risk factor, N (%) | 11421 (28.79%) | 4795 (26.75%) | 3497 (26.03%) | 4033 (34.57%) | 1630 (31.68%) | 555 (24.42%) | 298 (23.17%) |
| - 2 risk factors, N (%) | 12893 (32.5%) | 5947 (33.18%) | 4607 (34.3%) | 4051 (34.72%) | 1758 (34.16%) | 689 (30.31%) | 481 (37.4%) |
| - 3 or more risk factors, N (%) | 11269 (28.41%) | 5487 (30.61%) | 4398 (32.74%) | 2408 (20.64%) | 1251 (24.31%) | 886 (38.98%) | 430 (33.44%) |
| Cardiovascular diseases |  |  |  |  |  |  |  |
| - 0 CVD, N (%) | 8579 (21.63%) | 7294 (40.7%) | 2982 (22.2%) | 5454 (46.75%) | 2995 (58.2%) | 529 (23.27%) | 339 (26.36%) |
| - 1 CVD, N (%) | 6605 (16.65%) | 1772 (9.89%) | 1907 (14.2%) | 1017 (8.72%) | 464 (9.02%) | 365 (16.06%) | 176 (13.69%) |
| - 2 CVDs, N (%) | 14303 (36.06%) | 4814 (26.86%) | 4138 (30.8%) | 3032 (25.99%) | 917 (17.82%) | 729 (32.07%) | 412 (32.04%) |
| - 3 or more CVDs, N (%) | 10181 (25.67%) | 4043 (22.56%) | 4406 (32.8%) | 2164 (18.55%) | 770 (14.96%) | 650 (28.6%) | 359 (27.92%) |
| MMSE, mean (SD) | 28.05 (2.50) | 26.69 (3.66) | 27.93 (2.66) | 27.94 (2.83) | 26.81 (3.68) | 26.92 (3.56) | 25.66 (4.91) |
| MoCA, mean (SD) | 24.64 (4.28) | 20.51 (6.51) | 23.87 (4.96) | 22.38 (5.87) | 16.51 (7.15) | 23.07 (5.60) | 22.49 (6.10) |
| SAGEA, mean (SD) | 2.45 (2.85) | 2.53 (2.88) | 2.53 (2.87) | 1.87 (2.84) | 2.41 (2.98) | 3.21 (3.03) | 3.62 (3.82) |

N: number, IQR: interquartile range, BMI: body mass index, SD: standard deviation, CVD: cardiovascular disease (stroke, transient ischemic attack, myocardial infarction, heart failure, angina, atrial fibrillation, peripheral arterial disease), MMSE: Mini Mental State Examination, MoCA: Montreal Cognitive Assessment, SAGEA: Standardised Assessment of Global activities in the Elderly

**Supplementary Table 3.** Additional characteristics of participating individuals and countries in different world regions

|  | **North America, Oceania, Western Europe** | **South America** | **Eastern Europe**  **and Russia** | **East Asia** | **South Asia** | **Africa** | **West Asia** |
| --- | --- | --- | --- | --- | --- | --- | --- |
| Cardiovascular diseases |  |  |  |  |  |  |  |
| Stroke, N (%) | 4534 (11.43%) | 1029 (5.74%) | 1410 (10.5%) | 878 (7.53%) | 340 (6.61%) | 330 (14.52%) | 161 (12.52%) |
| Transient ischemic attack, N (%) | 1250 (3.15%) | 239 (1.33%) | 322 (2.4%) | 171 (1.47%) | 60 (1.17%) | 60 (2.64%) | 45 (3.5%) |
| Myocardial infarction, N (%) | 18092 (45.61%) | 6551 (36.55%) | 6169 (45.92%) | 3528 (30.24%) | 1243 (24.15%) | 900 (39.6%) | 557 (43.31%) |
| Heart failure, N (%) | 1715 (4.32%) | 1034 (5.77%) | 2298 (17.11%) | 767 (6.57%) | 184 (3.58%) | 93 (4.09%) | 49 (3.81%) |
| Angina (stable/unstable), N (%) | 13180 (38.38%) | 4356 (32.74%) | 4327 (37.92%) | 2899 (43.77%) | 955 (35.85%) | 771 (38.3%) | 432 (39.1%) |
| Atrial fibrillation, N (%) | 819 (2.06%) | 167 (0.93%) | 225 (1.67%) | 102 (0.87%) | 69 (1.34%) | 62 (2.73%) | 26 (2.02%) |
| Peripheral arterial disease, N (%) | 5180 (13.06%) | 2803 (15.64%) | 2185 (16.27%) | 878 (7.53%) | 518 (10.07%) | 384 (16.89%) | 133 (10.34%) |
| Cardiovascular risk factors |  |  |  |  |  |  |  |
| Hypertension, N (%) | 27948 (70.47%) | 12131 (67.69%) | 10077 (75.03%) | 7370 (63.18%) | 2870 (55.77%) | 1729 (76.07%) | 1004 (78.13%) |
| Diabetes mellitus, N (%) | 15177 (38.27%) | 7569 (42.23%) | 5441 (40.5%) | 3579 (30.68%) | 1488 (28.92%) | 1101 (48.44%) | 574 (44.67%) |
| Hyperlipidemia, N (%) | 5359 (13.51%) | 2901 (16.19%) | 1815 (13.51%) | 1054 (9.03%) | 394 (7.66%) | 476 (20.94%) | 232 (18.04%) |
| Non-cardiovascular diseases |  |  |  |  |  |  |  |
| Renal Dysfunction, N (%) | 730 (1.84%) | 606 (3.38%) | 360 (2.68%) | 177 (1.52%) | 236 (4.59%) | 77 (3.39%) | 25 (1.95%) |
| Liver disease, N (%) | 174 (1.13%) | 62 (0.9%) | 80 (1.34%) | 135 (3.06%) | 8 (0.89%) | 1 (0.16%) | 7 (1.93%) |
| Cancer, N (%) | 2988 (7.53%) | 834 (4.65%) | 508 (3.78%) | 443 (3.8%) | 98 (1.9%) | 67 (2.95%) | 58 (4.51%) |
| Fall, N (%) | 2751 (11.36%) | 1107 (10.03%) | 659 (8.84%) | 528 (7.27%) | 337 (7.93%) | 49 (2.98%) | 85 (9.21%) |
| Fracture, N (%) | 3871 (15.99%) | 1935 (17.54%) | 852 (11.43%) | 525 (7.23%) | 480 (11.3%) | 121 (7.36%) | 153 (16.58%) |
| Baseline BP and glucose measurements |  |  |  |  |  |  |  |
| Systolic BP, mmHg, mean (SD) | 139.53 (18.52) | 136.45 (19.27) | 138.51 (18.23) | 135.67 (16.58) | 132.01 (18.87) | 140.67 (20.22) | 139.8 (18.1) |
| Diastolic BP, mmHg, mean (SD) | 80.4 (10.84) | 79.12 (10.81) | 80.59 (10.7) | 79.11 (10.16) | 77.29 (10.64) | 82.17 (11.77) | 80.68 (10.92) |
| Fasting glucose, mmol/L, mean (SD) | 6.71 (2.42) | 6.51 (2.08) | 6.58 (2.25) | 6.23 (2.1) | 5.98 (1.96) | 6.98 (2.54) | 6.5 (2.39) |
| DCCT A1c %, mean (SD) | 6.47 (0.96) | 6.57 (0.94) | 6.51 (0.93) | 6.62 (1.04) | 6.36 (0.83) | 6.58 (1.03) | 6.61 (0.88) |
| Country income class |  |  |  |  |  |  |  |
| High, N (%) | 39668 (100%) | 1100 (6.14%) | 4609 (34.31%) | 4149 (35.56%) | 93 (1.81%) | 0 (0%) | 894 (69.52%) |
| Upper Middle, N (%) | 0 (0%) | 13842 (77.23%) | 4731 (35.22%) | 1577 (13.52%) | 601 (11.68%) | 841 (37%) | 39 (3.03%) |
| Lower Middle and Low, N (%) | 0 (0%) | 2981 (16.63%) | 4093 (30.47%) | 5941 (50.92%) | 4452 (86.51%) | 1432 (63%) | 353 (27.45%) |

N: number; BP: blood pressure; SD: standard deviation; DCCT: Diabetes Control and Complications Trial units

**Supplementary Table 4.** Unadjusted and adjusted differences in study-standardized baseline and first follow-up combined MMSE/ MoCA scores between different world regions, using the North America, Oceania and Western Europe group as reference.

|  | | **SA** | **EE+Russia** | **EAsia** | **SAsia** | **Africa** | **West Asia** |
| --- | --- | --- | --- | --- | --- | --- | --- |
| **Description** | **N** | **Mean Diff.(95%CI)** | **Mean Diff.(95%CI)** | **Mean Diff.(95%CI)** | **Mean Diff.(95%CI)** | **Mean Diff.(95%CI)** | **Mean Diff.(95%CI)** |
| Study standardized |  |  |  |  |  |  |  |
| **MoCA/ MMSE** |  |  |  |  |  |  |  |
| Unadjusted | 80777 | -0.533 (-0.591,-0.475) | -0.094 (-0.150,-0.038) | -0.135 (-0.198,-0.073) | -0.417 (-0.521,-0.313) | -0.223 (-0.361,-0.084) | -0.412 (-0.543,-0.281) |
| Age-sex adjusted | 80776 | -0.536 (-0.593,-0.479) | -0.139 (-0.195,-0.084) | -0.167 (-0.229,-0.105) | -0.493 (-0.596,-0.390) | -0.235 (-0.372,-0.098) | -0.462 (-0.591,-0.332) |
| Maximally-adjusted | 80126 | -0.452 (-0.508,-0.397) | -0.138 (-0.192,-0.085) | -0.113 (-0.173,-0.053) | -0.411 (-0.511,-0.311) | -0.203 (-0.335,-0.071) | -0.424 (-0.550,-0.299) |
| **SAGEA** |  |  |  |  |  |  |  |
| Unadjusted | 34317 | -0.071 (-0.165, 0.024) | -0.045 (-0.137, 0.046) | 0.304 (0.214, 0.393) | -0.217 (-0.436, 0.003) | -0.375 (-0.653, -0.097) | -0.448 (-0.680, -0.216) |
| Age-sex adjusted | 34317 | -0.067 (-0.162, 0.027) | -0.063 (-0.154, 0.028) | 0.283 (0.194, 0.372) | -0.241 (-0.460, -0.021) | -0.355 (-0.633, -0.078) | -0.480 (-0.711, -0.248) |
| Maximally adjusted | 33515 | 0.031 (-0.060, 0.122) | 0.032 (-0.056, 0.120) | 0.233 (0.146, 0.319) | -0.160 (-0.371, 0.050) | -0.320 (-0.585, -0.055) | -0.409 (-0.632, -0.186) |
| Country-standardized |  |  |  |  |  |  |  |
| **MoCA/ MMSE** |  |  |  |  |  |  |  |
| Unadjusted | 80777 | -0.094 (-0.149,-0.040) | -0.043 (-0.096, 0.010) | -0.075 (-0.135,-0.016) | -0.118 (-0.216,-0.019) | -0.006 (-0.137, 0.125) | -0.047 (-0.173, 0.078) |
| Age-sex adjusted | 80776 | -0.101 (-0.156,-0.047) | -0.094 (-0.147,-0.041) | -0.108 (-0.167,-0.050) | -0.200 (-0.297,-0.102) | -0.021 (-0.151, 0.108) | -0.098 (-0.223, 0.026) |
| Maximally-adjusted | 80126 | -0.016 (-0.069, 0.037) | -0.094 (-0.146,-0.042) | -0.057 (-0.115, 0.001) | -0.119 (-0.215,-0.023) | 0.012 (-0.115, 0.139) | -0.057 (-0.179, 0.065) |
| **SAGEA** |  |  |  |  |  |  |  |
| Unadjusted | 34317 | -0.066 (-0.152, 0.020) | -0.065 (-0.148, 0.018) | 0.022 (-0.060, 0.103) | -0.225 (-0.423,-0.027) | -0.154 (-0.406, 0.099) | -0.034 (-0.249, 0.181) |
| Age-sex adjusted | 34317 | -0.063 (-0.149, 0.023) | -0.085 (-0.168,-0.002) | 0.001 (-0.081, 0.082) | -0.251 (-0.449,-0.053) | -0.133 (-0.385, 0.119) | -0.067 (-0.282, 0.147) |
| Maximally adjusted | 33515 | 0.038 (-0.048, 0.123) | 0.006 (-0.077, 0.089) | -0.053 (-0.135, 0.028) | -0.178 (-0.374, 0.019) | -0.098 (-0.347, 0.151) | 0.003 (-0.209, 0.215) |

SA: South America; EE: Eastern Europe; EAsia: East Asia; SAsia: South Asia 95%CI: 95% confidence intervals, ICC: interclass correlation coefficient

Mini–Mental State Examination (MMSE) and Montral Cognitive Assessment (MoCA) are both based on 30-point questionnaires (scores 0-30); Standardized Assessment of Global activities in the Elderly (SAGEA) is based on a self-administered questionnaire (scores 0-45)

**Supplementary Table 5.** Logistic regression with absolute values used in the screening for cognitive or functional impairment by the respective study-standardized or country-standardized score thresholds.

|  | **Events/N(**≤**)** | **Events/N(>)** | **Cstatistic (95% CI)** | **Sensitivity** | **Specificity** | **Youden Index** | **Min. Distance** |
| --- | --- | --- | --- | --- | --- | --- | --- |
| *MMSE≤24 or MOCA≤25 for study standardized scores* | | | | | | | |
| ≤ -1.0 | 10814/ 10814 | 14213/ 69963 | 0.72 (0.71, 0.72) | 100% | 80% | 0.797 | 0.203 |
| ≤ -1.25 | 8125/ 8125 | 16902/ 72652 | 0.66 (0.66, 0.67) | 100% | 77% | 0.767 | 0.233 |
| ≤ -1.5 | 6901/ 6901 | 18126/ 73876 | 0.64 (0.64, 0.64) | 100% | 75% | 0.755 | 0.245 |
| *MMSE≤24 or MOCA≤25 for country standardized scores* | | | | | | | |
| ≤ -1.0 | 9818/ 11356 | 15209/ 69421 | 0.68 (0.68, 0.69) | 86% | 78% | 0.645 | 0.258 |
| ≤ -1.25 | 7855/ 8793 | 17172/ 71984 | 0.65 (0.65, 0.65) | 89% | 76% | 0.655 | 0.261 |
| ≤ -1.5 | 6048/ 6409 | 18979/ 74368 | 0.62 (0.61, 0.62) | 94% | 74% | 0.688 | 0.261 |
| *SAGEA≥7 for study standardized scores* | | | | | | | |
| ≤ -1.0 | 3700/ 5858 | 0/ 28459 | 0.96 (0.96, 0.97) | 63% | 100% | 0.632 | 0.368 |
| ≤ -1.25 | 3364/ 4301 | 336/ 30016 | 0.94 (0.93, 0.94) | 78% | 99% | 0.771 | 0.218 |
| ≤ -1.5 | 3105/ 3105 | 595/ 31212 | 0.92 (0.91, 0.93) | 100% | 98% | 0.981 | 0.019 |
| *SAGEA≥7 for country standardized scores* | | | | | | | |
| ≤ -1.0 | 3516/ 5169 | 184/ 29148 | 0.95 (0.94, 0.95) | 68% | 99% | 0.674 | 0.320 |
| ≤ -1.25 | 3293/ 4307 | 407/ 30010 | 0.93(0.92, 0.93) | 76% | 99% | 0.751 | 0.236 |
| ≤ -1.5 | 2867/ 3365 | 833/ 30952 | 0.88(0.87, 0.89) | 85% | 97% | 0.825 | 0.150 |

Mini–Mental State Examination (MMSE) and Montral Cognitive Assessment (MoCA) are both based on 30-point questionnaires (scores 0-30); Standardized Assessment of Global activities in the Elderly (SAGEA) is based on a self-administered questionnaire (scores 0-45)

**Supplementary Figure 1.** Fitting splines on the association between individual’s age cognitive or functional test scores. Box plot illustrates the median, interquartile range and range of scores in the whole study population.

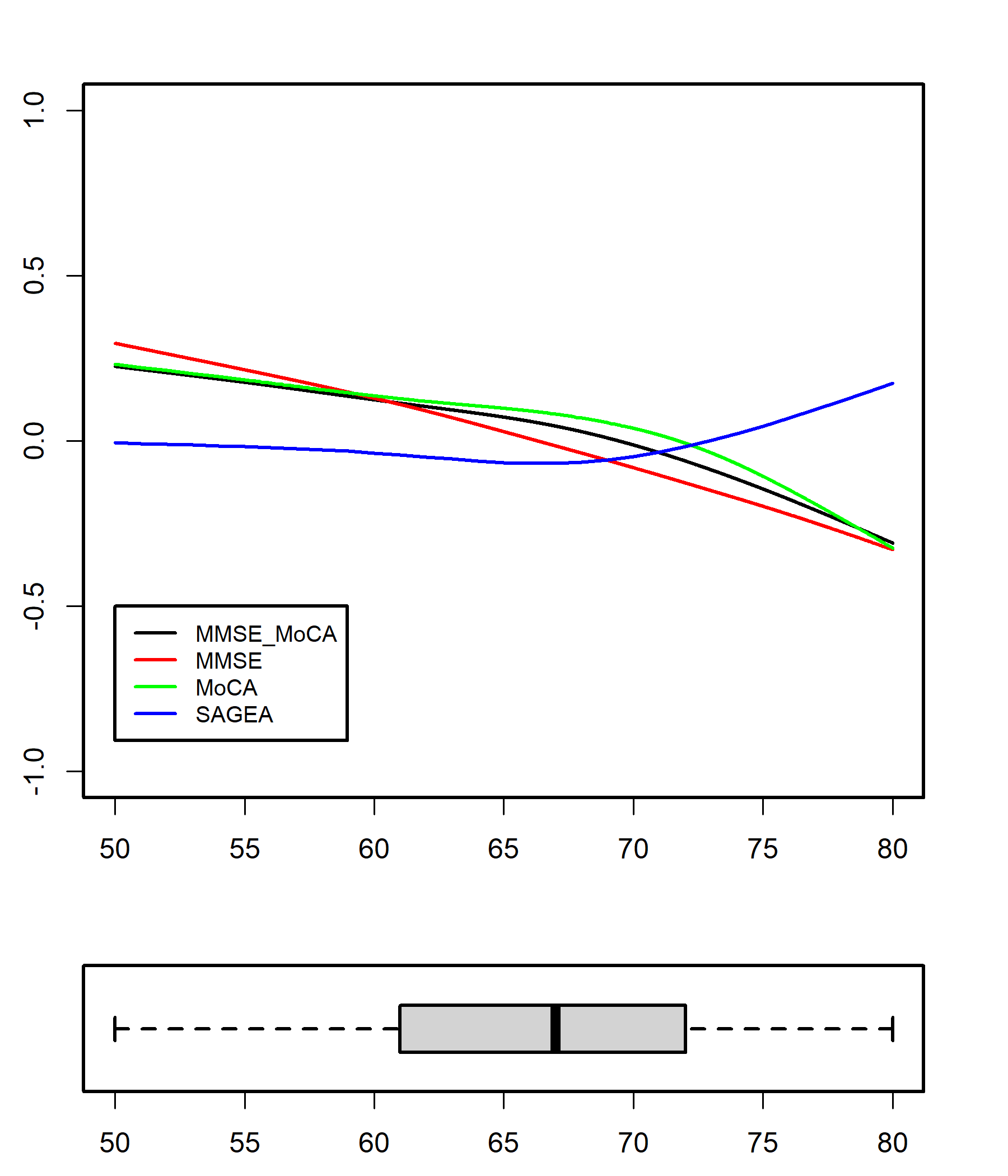

**Appendix.** Individual Items and Scoring for the Standard Assessment of Global Everyday Activities

**Over the past month, did you have any difficulties with the following:**

*Item scoring: none (0) or some -> if some, then mild (1), moderate (2), or severe (3)*

1. Keeping your attention or ‘train of thought’ during a conversation?
2. Remembering things that happened a few days before? (e.g., conversation, people visiting)
3. Ability to switch between things that are happening at the same time? (e.g., making tea and talking to someone)

**Over the past month, did you perform any of the following activities:**

*Item scoring: no (0) or yes -> if yes, difficulty? none (0) mild (1), moderate (2), or severe (3).*

4. Playing a game or reading a book that requires concentration (e.g., of games: crosswords, checkers, chess)
5. Finding your way around a new building? (e.g., hospital/clinic)
6. Organizing a trip or social activities? (e.g., vacation or family occasion) (score the activity that the person finds to be the more difficult of the two)
7. Doing your own finances or shopping? (score the activity that the person finds to be the more difficult of the two)
8. Organizing and taking your medications?
9. Preparing a meal and/or doing laundry? (score the activity that the person finds to be more difficult of the two)
10. a) Driving? Do not drive (go to 10b)

10. b) Using public transportation? Do not use (go to 11)

**Over the past month, did you perform any of the following activities:**

*Item scoring: no (0) or yes -> if yes, difficulty? none (0) mild (1), moderate (2), or severe (3). If requires help, add 1 point to maximum score of 3 points for that item. If person did not do item 12, score 3 points.*

11. Using stairs? (one flight) If yes, did you require help?
12. Walking? (about 10m or 32ft or 14 steps) If yes, did you require help?

**Over the past month, did you perform any of the following activities:**

*Item scoring: no (3) or yes -> if yes, difficulty? none (0) mild (1), moderate (2), or severe (3). If requires help, add 1 point to a maximum score of 3 points for that item. If person did not do activity, score 3 points.*

13. Dressing? If yes, did you require help?
14. Transferring from bed to chair? If yes, did you require help?
15. Bathing or toileting? (score the activity that the person finds to be more difficult of the two)
